# Dual T-cell constant β chain (TRBC)1 and TRBC2 staining for the identification of T-cell neoplasms by flow cytometry

**DOI:** 10.1101/2023.12.01.23299254

**Authors:** Pedro Horna, Matthew J Weybright, Mathieu Ferrari, Dennis Jungherz, YaYi Peng, Zulaikha Akbar, F Tudor Ilca, Gregory E Otteson, Jansen N Seheult, Janosch Ortmann, Min Shi, Paul M Maciocia, Marco Herling, Martin A Pule, Horatiu Olteanu

**Author notes:** **Correspondence:** Pedro Horna, M.D. Mayo Clinic, 200 First Steet SW, Rochester, MN 55905. **COMPETING INTERESTS STATEMENT**: MP and PMM are inventors on a patent describing use of TRBC1/2 for diagnosis and treatment of T cell malignancies. MP and MF are inventors on a patent describing TRBC2 antibodies. Autolus Therapeutics owns patents claiming use of TRBC1/2 for diagnosis. MP, MF and ZA own stock in and are employees of Autolus Therapeutics. FTI is a former Autolus employee. All other authors report no conflicts of interest to disclose.

## Abstract

The diagnosis of leukemic T-cell malignancies is often challenging, due to overlapping features with reactive T-cells and limitations of currently available T-cell clonality assays. Recently developed therapeutic antibodies specific for the mutually exclusive T-cell receptor constant β chain (TRBC)1 and TRBC2 isoforms provide a unique opportunity to assess for TRBC-restriction as a surrogate of clonality in the flow cytometric analysis of T-cell neoplasms. To demonstrate the diagnostic utility of this approach, we studied 135 clinical specimens with (50) or without (85) T-cell neoplasia, in addition to 29 blood samples from healthy donors. Dual TRBC1 and TRBC2 expression was studied within a comprehensive T-cell panel, in a fashion similar to the routine evaluation of kappa and lambda immunoglobulin light chains for the detection of clonal B-cells. Polytypic TRBC expression was demonstrated on total, CD4^+^ and CD8^+^ T-cells from all healthy donors; and by intracellular staining on benign T-cell precursors. All neoplastic T-cells were TRBC-restricted, except for 5 cases (10%) lacking TRBC expression. T-cell clones of uncertain significance were identified in 15 samples without T-cell malignancy (13%), and accounted for smaller subsets than neoplastic clones (median: 4.7% vs. 73% of lymphocytes, p<0.0001). Single staining for TRBC1 produced spurious TRBC1-dim subsets in 21 clinical specimens (16%), all of which resolved with dual TRBC1/2 staining. Assessment of TRBC restriction by flow cytometry provides a rapid diagnostic method to detect clonal T-cells, and to accurately determine the targetable TRBC isoform expressed by T-cell malignancies.

## INTRODUCTION

The laboratory diagnosis of leukemic T-cell lymphoproliferative disorders neoplasms relies on the identification of cytologically and/or immunophenotypically abnormal T-cell populations, often in correlation with clinical findings and ancillary laboratory testing.^1^ However, confident interpretation of an atypical T-cell population as diagnostic for T-cell malignancy is often challenging, given that similar atypical T-cell subsets can occasionally be encountered in reactive settings. Moreover, some leukemic T-cell neoplasms may lack overt immunophenotypic aberrancies and/or convincing cytologic abnormalities, precluding an unequivocal laboratory diagnosis.^2,3^

The diagnosis of B-cell neoplasms has been largely facilitated by the broad utilization of stains for kappa and lambda immunoglobulin light chains to assess for immune receptor monotypia indicative of B-cell clonality.^4^ Such a simplified and effective approach has not been available for the analysis of T-cells in routine clinical diagnostics. Instead, detection of clonal T-cell receptor (TCR) gene rearrangements by multiplex polymerase chain reaction is commonly utilized^5^. This is a qualitative approach that lacks immunophenotypic information, is subject to interpretative challenges^6^, and can produce false positive results in the setting of aging and inflammation.^7,8^ Alternatively, analysis of the TCR variable β repertoire (TCR-Vβ) by flow cytometry can be used,^9,10^ but this method is labor-intensive, costly, often difficult to interpret and of limited sensitivity.^11^

The gene encoding the TCR constant β chain (TRBC) has two isoforms: TRBC1 and TRBC2. In a manner analogous to light chain restriction, TCR gene rearrangement results in a TCR with the TCR β chain constant region encoded by either TRBC1 or TRBC2. This aspect of TCR gene-rearrangement has been described decades ago but is often overlooked, likely because unlike κ and λ light chains, TRBC1/2 encode almost identical proteins. We previously demonstrated that the anti-TCR antibody (JOVI.1)^12^ has exquisite sensitivity for TRBC1, and this selectivity can be exploited to develop novel CAR T-cell therapies to deplete TRBC1^+^ malignant and benign T-cells while preserving T-cell immunity maintained by the TRBC2^+^ immune repretoire.^13^ More recently, we employed computational biology and protein engineering to rationally design and produce mutant versions of JOVI.1 with switched specificity for TRBC2 and pre-clinical activity as CAR T-cell constructs targeting TRBC2^+^ T-cell malignancies.^14^

The availability of complementary antibodies against TRBC1 and TRBC2 provides a unique opportunity to easily demonstrate T-cell clonality, in a fashion similar to detecting clonal B-cells based on kappa or lambda immunoglobulin light chain restriction. We hereby demonstrate the optimal performance of this approach for the confident laboratory diagnosis of T-cell neoplasms. In addition, we show that dual assessment of TRBC1/TRBC2 expression eliminates spurious TRBC-dim subsets on previously described TRBC1-only staining approaches, allowing for the accurate determination the targetable TRBC isoform expressed by T-cell malignancies.

## MATERIALS AND METHODS

### Anti-TRBC2 antibody development

A mouse anti-human TRBC2 antibody was specifically designed and developed for diagnostic flow cytometry, following a previously described strategy.^14^ In short, an anti-TRBC2 antibody was produced based on structural engineering and rational design of mutations on complementarity determining region (CDR)1 (T28K and Y32F) and CDR3 (A96N and N99M) of the JOVI.1 antibody (Kabat numbering scheme), resulting in switched antibody specificity from TRBC1 to TRBC2 (**Figure 1A**). The kinetic profile of the anti-TRBC2 antibody was studied against soluble TRBC1^+^ or TRBC2^+^ TCRs on a Biacore T200 surface plasmon resonance system (Cytiva, Marlborough, MA); and its thermal stability was assessed on a via Prometheus NT.48 nanoDSF differential scanning fluorimeter (NanoTemper, München, Germany) (**Supplemental Figure 1**). Wild-type (WT) Jurkat cells (TRBC1^+^) and Jurkat cells engineered to express TRBC2 were evaluated by flow cytometry to confirm similar levels of surface CD3/TCR expression (**Supplemental Figure 2**). These cell lines were then utilized to study the specificity and affinity of the anti-TRBC2 antibody compared to JOVI.1, using a secondary anti-mouse IgG (H+L) AF647 antibody (Invitrogen, Waltham, MA), and staining with anti-CD3-PECy7 (Biolegend, San Diego, CA) to gate on surface CD3/TCR-positive cells.

**FIGURE 1.**
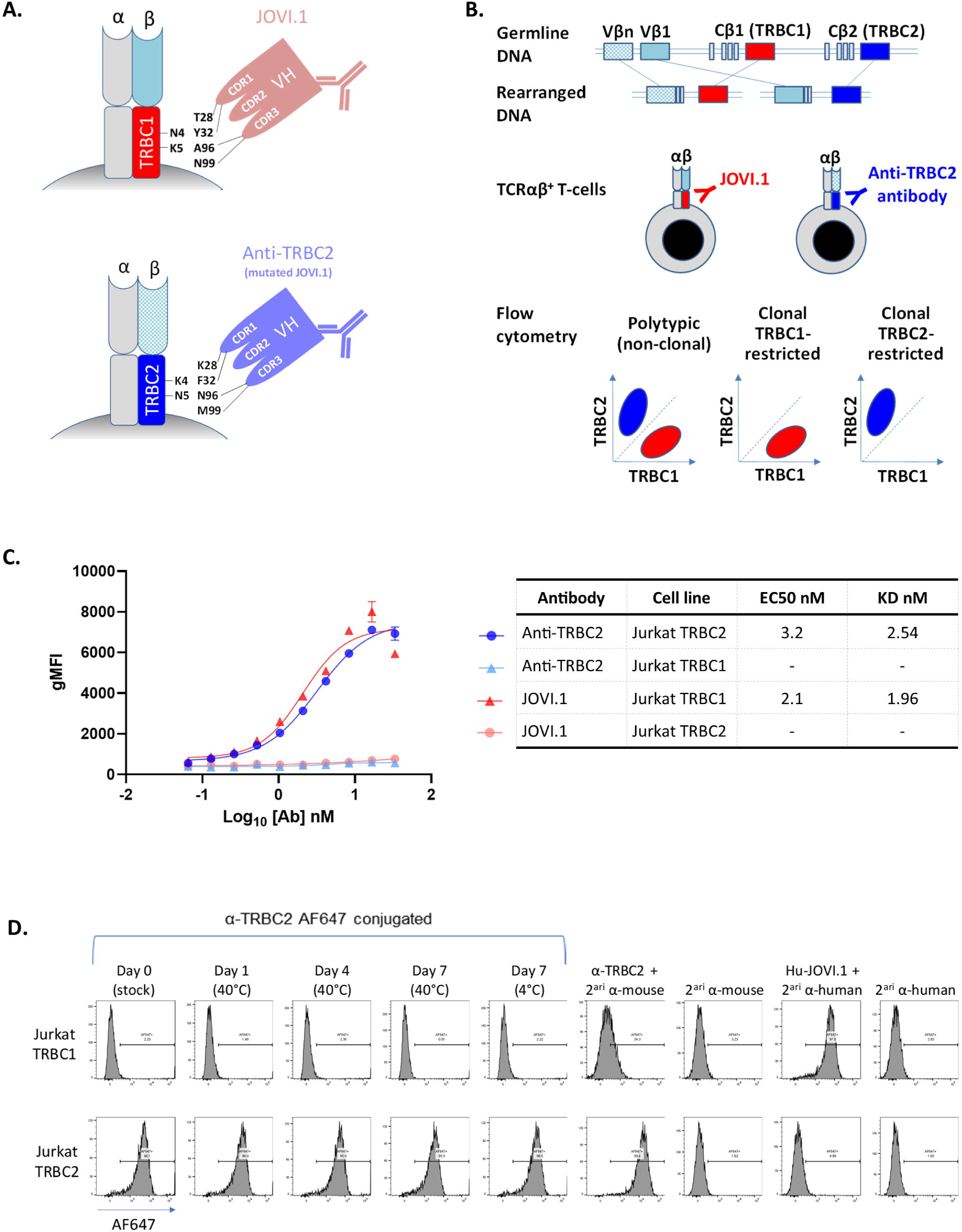
Strategic mutations on complementarity determining regions (CDR) of the JOVI.1 antibody result in switched specificity from T-cell receptor constant β chain (TRBC)1 to TRBC2, allowing for dual TRBC staining by flow cytometry. **A.** Simplified model of JOVI.1 antibody binding to TRBC1 (top), showing key amino-acid residues on both molecules responsible for the discriminative binding to one isoform only. Rationally-designed antibody mutations on CDR1 (T28K and Y32F) and CDR3 (A96N and N99M) of JOVI.1’s variable heavy chain (VH) domain result in switched specificity to TRBC2 (bottom). **B.** T-cell receptor αβ gene rearrangement showing the random selection of 1 of 2 mutually exclusive TRBC genes. Anti-TRBC1 (JOVI.1) and anti-TRBC2 antibodies can be utilized in conjunction to assess for TRBC-restriction by flow cytometry as a surrogate for T-cell clonality. **C.** Half maximal effective concentration (EC50) measurements and dissociation constant (KD) estimations (non-linear regression one site binding analysis) of anti-TRBC2 (blue and light blue) or JOVI.1 (red and pink) binding to TRBC1-positive (triangles) or TRBC1-posiitve (circles) Jurkat cells; as assessed by geometric mean fluorescence intensity (gMFI) using an anti-mouse IgG fluorescent-labelled secondary antibody. **D.** Flow cytometry histograms showing specificity of the anti-TRBC2 antibody for TRBC2-positive (top) as compared to TRBC1-positive (bottom) Jurkat cells; and stability of an Alexa Fluor(AF)-647-conjugated anti-TRBC2 antibody. Secondary (2^ari^) antibodies were AF647 conjugates. A humanized JOVI.1 antibody (Hu-JOVI.1) is also shown as control.

### Clinical Specimens

As part of a test development effort, fresh clinical samples received for flow cytometric analysis at Mayo Clinic, Rochester, MN, were selected based on the presence or absence of involvement by a T-cell lymphoproliferative disorder, and availability of additional material remaining after diagnostic work up (**Table 1**). Peripheral blood specimens from healthy donors were obtained from Mayo Clinic’s biospecimen program. All test development data, diagnostic laboratory data, and corresponding medical records were retrospectively reviewed. This study was approved by the Mayo Clinic Institutional Review Board.

### Flow cytometry panels

Two comprehensive 11-color single-tube panels (“T-cell” and “Sezary” panels), including anti-TRBC1 and anti-TRBC2 antibodies, were developed at Mayo Clinic, Rochester, MN. In addition, a TCR-Vβ repertoire flow cytometry kit (IOTest Beta Mark, Beckman Coulter, Brea, CA) was modified to assess for TRBC1 and TRBC2 expression in combination with TCR-Vβ classes on selected healthy donors (“Vβ/TRBC” panel). On all panels, our anti-TRBC2 antibody was pre-conjugated to Alexa Fluor 647. The T-cell and Sezary panels were designed with a FITC-conjugated JOVI.1 (anti-TRBC1) antibody (Ancell Corporation, Bayport, MN, USA), while the Vβ/TRBC panel included a Brilliant Violet 605-conjugated JOVI.1 antibody (BD Biosciences, Franklin Lakes, NJ) (**Table 2**). All surface staining steps were performed as previously described.^15^ In cases where the cells of interest lacked surface expression of CD3 or TRBC, staining for cytoplasmic TRBC1 and TRBC2 (with or without cytoplasmic CD3) was performed after surface staining for all other antigens and standard fixation and permeabilization (FIX & PERM, Invitrogen, Waltham, MA). Events were acquired on a FACSLyric flow cytometer (BD Biosciences), with a target of 100,000 total cells. Routine diagnostic flow cytometry outside this test development activity was performed on clinical specimens using our TRBC1-only T-cell^15^ and TRBC-1 only Sezary^16^ panels, as previously described.

### Flow cytometry data analysis

Common normal T-cell subsets and immunophenotypically abnormal subsets consistent with a neoplastic T-cell population were manually gated by expert hematopathologists (PH and HO) on Kaluza version 2.1 (Beckman Coulter), based on patterns of expression of common surface T-cell antigens excluding TRBC1 and TRBC2. Expression of TRBC1 and TRBC2 on gated T-cell subsets of at least 200 events was evaluated on a TRBC1 vs. TRBC2 dot plot to assess for TRBC-restriction as surrogate for clonality (**Figure 1B**). Thresholds for clonality were arbitrarily defined based on our extensive experience assessing TRBC1 expression only on clinical specimens (<15% or >85% TRBC1-positive events),^15–19^ and applied on this study as percentage positivity (>85% TRBC1- or TRBC2-positive events) or equivalent TRBC2:TRBC1 ratios (TRBC2:TRBC1 <0.18 or >5.7). The development data using TRBC1/TRBC2 dual staining was compared to data collected during routine flow cytometric analysis using our previously validated TRBC1-only panels.^15,16^

### Statistical analysis

All statistic calculations were performed using GraphPad Prism, version 10.0.2 for Windows (GraphPad Software, San Diego, CA, USA). Comparisons of measurement values between two groups were performed using the Mann Whitney test (clone size and TRBC2:TRBC1 ratios) or an unpaired 2-tailed t test (TCR-Vβ class percentages). A receiver operating characteristic (ROC) curve was constructed based on the maximum of %TRBC1^+^ and %TRBC2^+^ events for each gated TRBC-expressing tumor population (true positives); compared to a similar maximum for each gated CD4^+^/CD8^-^, CD8^+^/CD4^-^, CD4^+^/CD8^+^ and CD4^-^/CD8^-^ TCRαβ T-cell subset (>200 events) from samples without T-cell neoplasia (true negatives). A statistically significant P value was considered as less than 0.05.

## RESULTS

### A novel anti-TRBC2 antibody combined with JOVI.1 demonstrates TRBC polytypia on benign blood and bone marrow T-cells by flow cytometry

We first tested the specificity of a newly developed and strategically mutated JOVI.1 antibody (**Figures 1A and 1B**) with a kinetic profile favoring recognition of TRBC2 over TRBC1 (**Supplemental Figure 1A**). By flow cytometry, this anti-TRBC2 antibody bound to the surface of a genetically engineered TRBC2-positive Jurkat cell line, but not to wild type TRBC1-positive Jurkat cells exhibiting comparable levels of CD3/TCR expression (**Figure 1C** **and Supplemental Figure 2**). Moreover, the cell line-based binding affinity for TRBC2 was comparable to that of JOVI.1 for TRBC1, based on similar dissociation constants (KD of 2.54 nM vs. 1.96nM, respectively). Labelling of anti-TRBC2 with AF647 resulted in a thermally stable flow cytometry reagent (**Supplemental Figure 1B**) with preserved specific staining of TRBC2^+^ Jurkat cells only (**Figure 1D**).

We then studied peripheral blood specimens from 25 healthy donors, using a single-tube flow cytometry T-cell panel, including anti-TRBC2, JOVI.1 (anti-TRBC1), TCRγδ (to gate on TCRαβ by exclusion) and other antibodies recognizing common T-cell antigens (**Tables 1 and 2**). In all specimens, gated total TCRαβ T-cells, CD4^+^ T-cells and CD8^+^ T-cells showed TRBC2:TRBC1 ratios consistent with polytypia, as evaluated using clonality thresholds based on our extensive experience using JOVI.1 staining only^15,16,18–20^ (**Figures 2A and 2B**). These clonality thresholds were further validated on receiver operating characteristic (ROC) curve analysis using tumor and controls samples from this study (**Supplemental Figure 3**). On healthy donors, the median TRBC2:TRBC1 ratios and interquartile ranges were 1.6 (1.3-2.0) for total TCRαβ T-cells, 1.3 (1.2-1.6) for CD4^+^ T-cells, and 1.8 (1.5-2.7) for TCRαβ CD8^+^ T-cells; with slightly higher TRBC2:TRBC1 ratios for CD8^+^ T-cells compared to CD4^+^ T-cells (p<0.0001).

**FIGURE 2.**
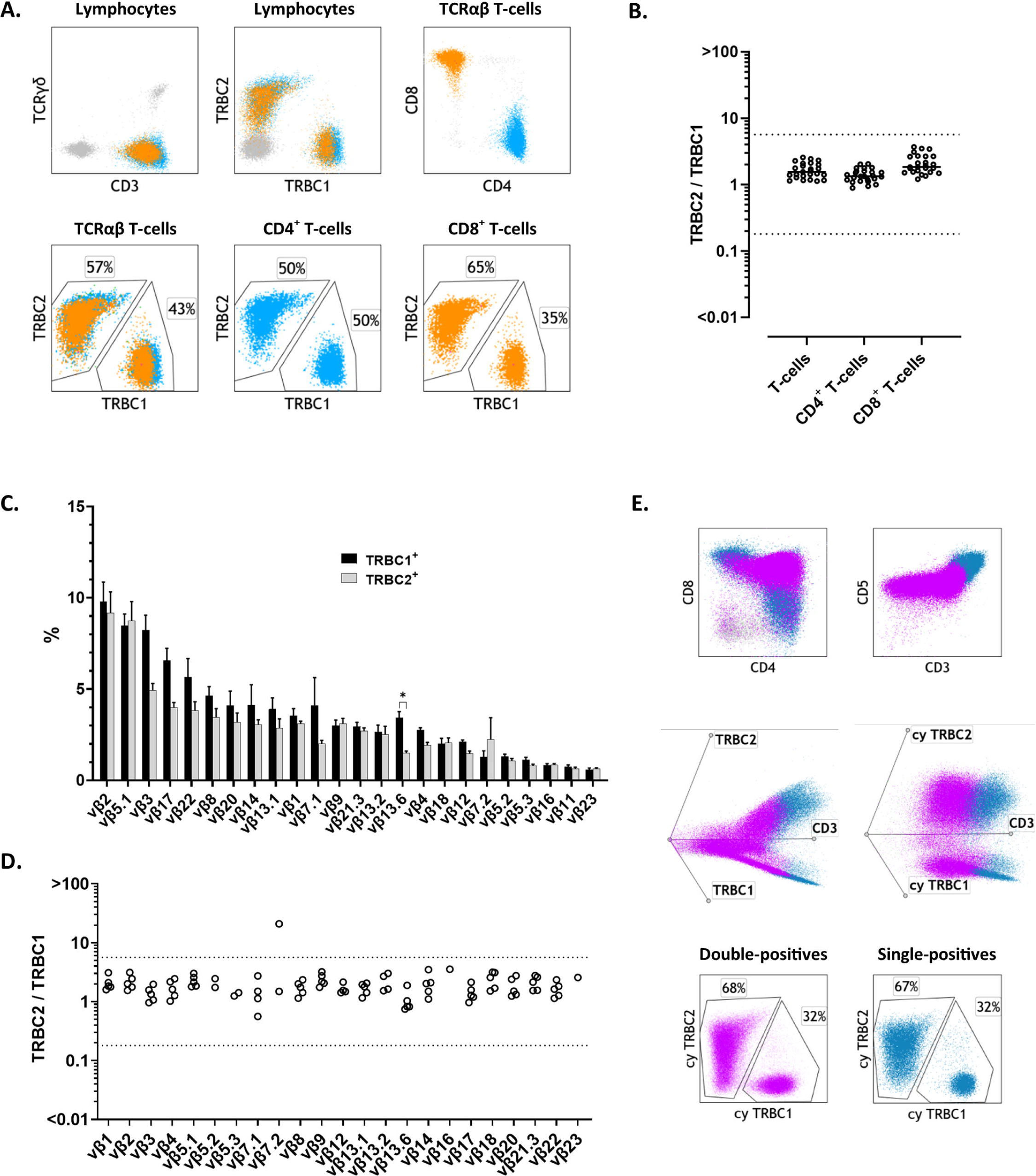
Dual staining for T-cell receptor constant β chain (TRBC)1 and TRBC2 demonstrates mutually exclusive and polytypic TRBC expression on benign T-cells, T-cell precursors, and T-cell receptor variable β (TCR-Vβ) subsets. **A.** Representative peripheral blood flow cytometry findings on a healthy donor showing polytypic TRBC expression on total, CD4^+^ (cyan) and CD8^+^ (orange) T-cells. **B.** TRBC2:TRBC1 ratios of total, CD4^+^ and CD8^+^ T-cells from 25 healthy donor’s peripheral blood specimens. Solid lines: medians. Dotted lines: thresholds for clonality. **C.** TCR-Vβ repertoire by flow cytometry on gated TRBC1^+^ (black bars) and TRBC1^-^ (grey bars) peripheral blood T-cells from 5 healthy donors, showing remarkably similar distributions. (*): p<0.05. **D.** TRBC2:TRBC1 ratios of each TCR-Vβ-positive T-cell subset from 5 healthy donors. Dotted lines depict thresholds for clonality. **E.** Representative flow cytometry findings of benign maturing T-cell precursors on a mediastinal mass involved by thymoma. CD4/CD8-double-positive T-cell precursors (*violet*) gradually acquire surface CD3 expression in addition to surface TRBC1 and TRBC2 expression, before maturing into overtly polytypic single-positive T-cells (*blue*). Cytoplasmic (cy) TRBC1 and TRBC2 staining demonstrates polytypic double positive (*violet*) and single positive (*blue*) T-cell precursors.

### TRBC1 and TRBC2 expression by flow cytometry is independent of TCR-Vβ restriction and detectable in the cytoplasm of maturing T-cell precursors

TCR-Vβ gene usage and TRBC gene selection are theoretically independent and probabilistically unrelated events during T-cell receptor gene rearrangement. This is a key assumption in the simplification of clonality assessment using TRBC1 and TRBC2 staining, as preferential TRBC usage associated with a subset of Vβ genes would produce false positive clonality results in settings where these Vβ subsets are enriched. To confirm this requirement, we compared the TCR-Vβ repertoire of gated TRBC1^+^ and TRBC2^+^ peripheral blood T-cells by flow cytometry on 5 healthy donors. As anticipated, both TCR-Vβ repertoires closely mirrored each other, with no discernable bias except for a slight but statistically significant skew of TCR-Vβ 13.6 for TRBC1 over TRBC2 (median: 3.4% vs. 1.5%; adjusted p=0.01, Holm-Sidak method) (**Figure 2C**). We also directly calculated the TRBC2:TRBC1 ratio of each TCR-Vβ subset on these 5 healthy donors. All detectable Vβ-positive subsets (≥200 cells) on all studied patients showed a polytypic TRBC2:TRBC1 ratio (**Figure 2D**), except for a single donor harboring a small subset of TRBC2-restricted Vβ7.2^+^ T-cells (4.1% of lymphocytes) corresponding to a small CD8^+^ T-cell clone of uncertain significance (T-CUS)^15^ on further evaluation (**Supplemental Figure 4**).

As TCRβ gene rearrangement and protein expression are known to occur during early T-cell development^21^, we hypothesized that cytoplasmic staining for TRBC1 and TRBC2 by flow cytometry should be able to demonstrate TRBC polytypia on benign immature T-cell precursors, despite the negative to dim surface TCR expression on most cells. To confirm this, we evaluated surface and cytoplasmic TRBC1 and TRBC2 expression (T-cell panel) on 5 surgically excised thymic masses harboring benign immature T-cell precursors in the setting of thymic epithelial tumors. In all cases, CD4^+^/CD8^+^ double-positive and single-positive T-cell precursors showed detectable expression of cytoplasmic TRBC1 and TRBC2 by flow cytometry, with a polytypic TRBC2:TRBC1 ratio discernable throughout a broad maturation spectrum of progressive surface CD3/TCR expression (**Figure 2E**).

### TRBC-restriction is characteristic of T-cell neoplasms and small T-cell clones of uncertain significance

We next evaluated 50 clinical specimens with confirmed involvement by various TCRαβ T-cell neoplasias, including cutaneous T-cell lymphoma (CTCL, 21), T lymphoblastic leukemia/lymphoma (T-LBL, 10), T-cell large granular lymphocytic leukemia (T-LGLL, 7), peripheral T-cell lymphoma (PTCL, 6), T-cell prolymphocytic leukemia (T-PLL, 3), lymphocytic variant of hypereosinophilic syndrome (T-HES, 2), and hepatosplenic T-cell lymhpoma (HSTCL, 1) (**Tables 1 and 2**). In 14 cases (28%, 7 T-LBL, 4 CTCL, 1 PTCL, 1 HSTCL and 1 L-HES) cytoplasmic TRBC1 and TRBC2 staining was performed to assess for TRBC-restriction in the setting of dim to negative surface CD3 and/or TRBC expression. In all but 5 cases, expert manual analysis demonstrated a clonal TRBC2:TRBC1 ratio indicative of T-cell clonality on the gated neoplastic cells (**Figures 3A-3D**). The remainder 5 neoplastic cases (10%) showed aberrant loss of surface and cytoplasmic TRBC expression on the tumor cells (3 T-ALL, 1 CTCL and 1 L-HES; **Figure 3E**). Both TRBC1-restricted and TRBC2-restricted tumors were observed in all disease categories studied, with an overall higher incidence of TRBC2^+^ neoplasms (76%, 95% CI: 62-87%) as expected by the slight overrepresentation of TRBC2 in the normal T-cell repertoire. Moreover, all background total T-cells, CD4^+^ T-cells and CD8^+^ T-cells detectable (≥200 events) outside the expert-defined neoplastic gate showed a polytypic TRBC2:TRBC1 ratio, demonstrating that TRBC-restriction is a feature specific for the neoplastic T-cell subset (**Figure 4A**).

**FIGURE 3.**
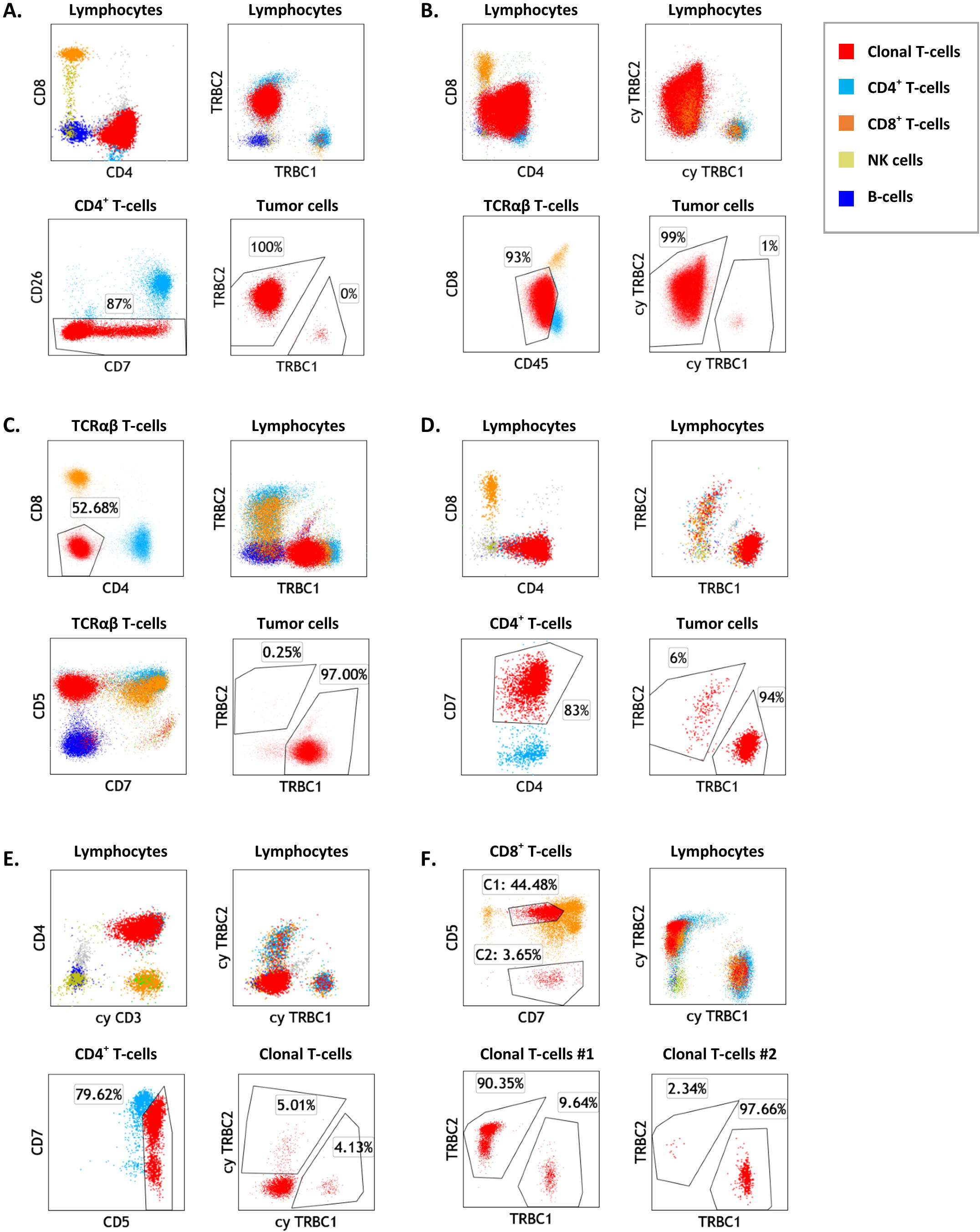
Dual T-cell receptor constant β chain (TRBC)1 and TRBC2 staining by flow cytometry demonstrates TRBC restriction on gated malignant T-cells. **A.** Representative flow cytometry plots of peripheral blood involvement by Sezary syndrome (Sezary panel), showing a distinctly abnormal CD4^+^ T-cell subset (red) with loss of CD26 expression and TRBC2-restriction. Also shown are background polytypic CD4^+^ T-cells (cyan), CD8^+^ T-cells (orange), NK cells (gold) and B-cells (blue). **B.** Peripheral blood involvement by T lymphoblastic leukemia/lymphoma (T-cell panel), showing an abnormal CD4-variable/CD8-dim T-cell population (red) that was surface CD3/TCR negative (data not shown) and on which TRBC2-restriction could be demonstrated by cytoplasmic (cy) TRBC1 and TRBC2 staining. **C.** Inguinal lymph node involvement by cutaneous T-cell lymphoma, not otherwise specified, showing an expanded CD4/CD8 double-negative T-cell subset with TRBC1-restriction. **D.** Cervical lymph node biopsy involved by a CD4-positive peripheral T-cell lymphoma, showing a large subset of CD4^+^/CD7^+^ T-cells with TRBC1-restriction and absence of overt immunophenotypic aberrancies. **E.** Peripheral blood from a patient with lymphocytic variant of hypereosinophilic syndrome, showing an abnormal CD4^+^ T-cell subset negative for surface CD3 (not shown), positive for cytoplasmic CD3, and negative for surface and cytoplasmic TRBC. **F.** Peripheral blood from a patient with Felty syndrome, showing 2 small CD8^+^ T-cell subsets with opposite TRBC-restriction, consistent with T-cell clones of uncertain significance.

**FIGURE 4.**
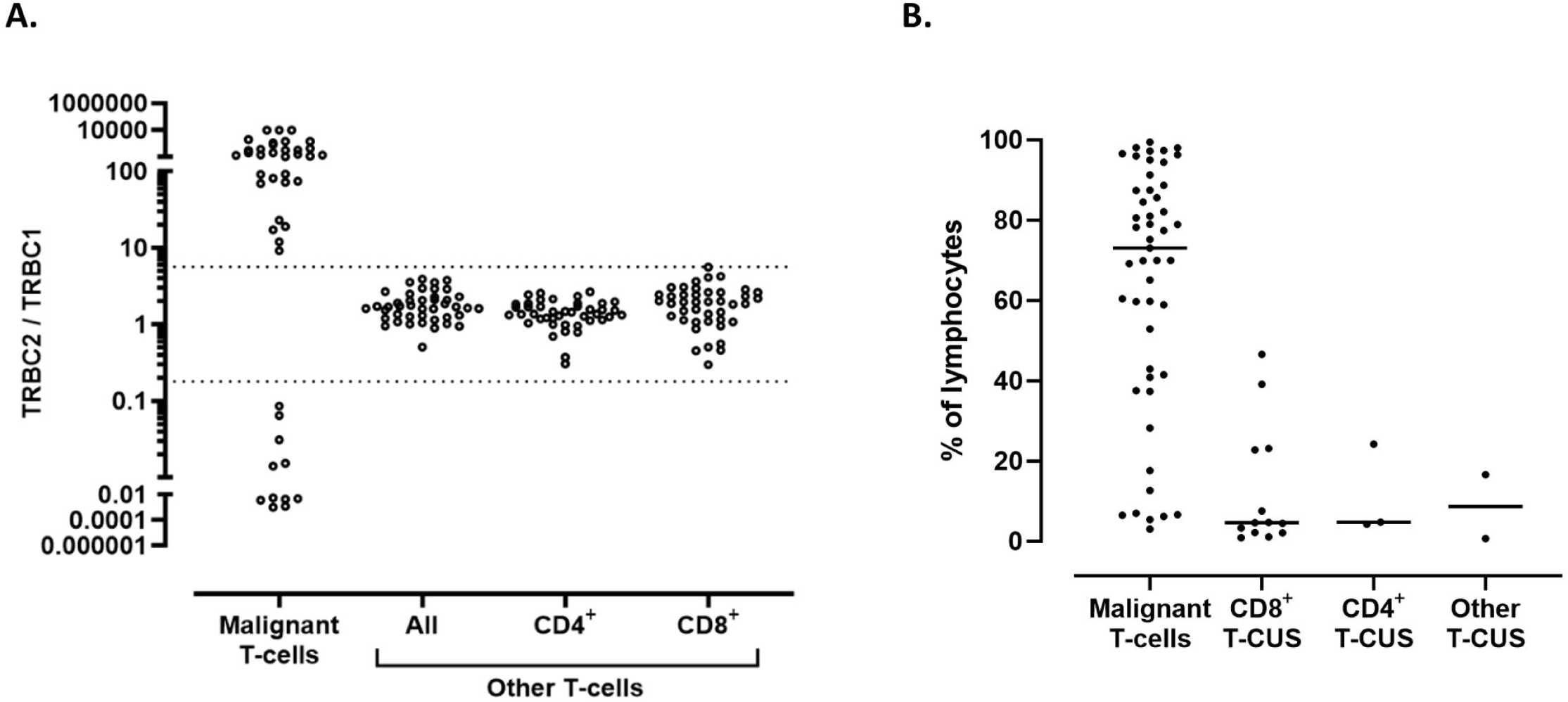
Evaluation of T-cell receptor constant β chain (TRBC)2:TRBC1 ratio on T-cell subsets identifies malignant populations and small T-cell clones of uncertain significance (T-CUS). **A.** TRBC2:TRBC1 ratios of gated malignant T-cell populations from 45 clinical specimens with various T-cell neoplasms (excluding 5 neoplasms lacking intracellular TRBC expression). Also shown are TRBC2:TRBC1 ratios of background (non-malignant) T-cells and major T-cell subsets. Dotted lines: thresholds for clonality. **B.** Clone size of 50 T-cell neoplasms expressed as percentage of lymphocytes, compared to 18 T-CUS detected on 15 benign specimens.

Using a similar expert analysis, we evaluated 85 clinical specimens from patients with no T-cell malignancy and 29 blood specimens from healthy donors, for T-cell subsets with TRBC1 or TRBC2 restriction. Overall, 18 immunophenotypically distinct TRBC-restricted subsets were identified on 5 (17%) donors and 10 (12%) patients with no T-cell malignancy (**Figure 3F**). The features of these small subsets were similar to those previously described for T-cell clones of uncertain significance,^15^ including mostly CD8^+^ phenotype (72%), lack of overt tumor-specific immunophenotypic abnormalities, and much smaller median clone size than malignant T-cell subsets (median: 4.7% vs. 73% of lymphocytes, p<0.0001, **Figure 4B**).

## DISCUSSION

We hereby introduce a novel strategy to identify TCRαβ T-cell neoplasms by flow cytometry, based on the restricted expression of TRBC1 or TRBC2. This approach resembles the routine and broadly utilized assessment of kappa and lambda immunoglobulin light chain restriction for identification of B-cell malignancies^4^ (**Figure 1B**). It also relies on expert manual segregation (gating) of T-cell subsets exhibiting distinct immunophenotypic features or increased relative abundance concerning for a neoplastic process (**Figure 3, A-D**), currently the standard of care in clinical flow cytometric analysis of T-cells.^1,2,22–24^ Rapid confirmation of TRBC-restriction on gated atypical T-cell subsets using TRBC1 and TRBC2 stains within the same panel greatly assists the interpretation of flow cytometry results, obviating the need for a separate T-cell clonality testing in most settings. Given the relative ease by which this novel strategy can be implemented in routine clinical practice, the significant added value to the interpretation of immunophenotypically suspicious T-cell subsets and the potential cost savings, we believe that dual TRBC1/TRBC2 staining is likely to be adopted as a new laboratory standard in the clinical flow cytometric evaluation of leukemic T-cell neoplasms.

Our proposed dual TRBC1 and TRBC2 staining strategy is built upon our extensive experience using TRBC1-only staining in clinical diagnostics^15–20,25,26^, in addition to similar reports from few other laboratories.^27–29^ In this study, single staining for TRBC1 using our previously validated diagnostic panels produced spurious TRBC1-dim subsets in 21 (16%) clinical specimens, all of which completely resolved using dual TRBC1/TRBC2 staining in our experimental panels (**Figure 5**). These artifacts are reminiscent of our historical experience using only kappa or only lambda immunoglobulin light chain staining with 3 to 4 color legacy flow cytometry panels, a practice that is now obsolete. Dual staining for TRBC1 and TRBC2 significantly improves the confident and reproducible separation of TRBC1^+^ and TRBC2^+^ T-cell events, a critical step in assessing T-cell clonality based on TRBC restriction. As such, we anticipate that dual assessment of TRBC1 and TRBC2 expression on 2-dimensional plots (as routinely performed for kappa and lambda immunoglobulin light chains) will be strongly favored, effectively overcoming common technical artifacts and providing and accurate determination of the targetable TRBC isoform expressed by T-cell malignancies.

**FIGURE 5.**
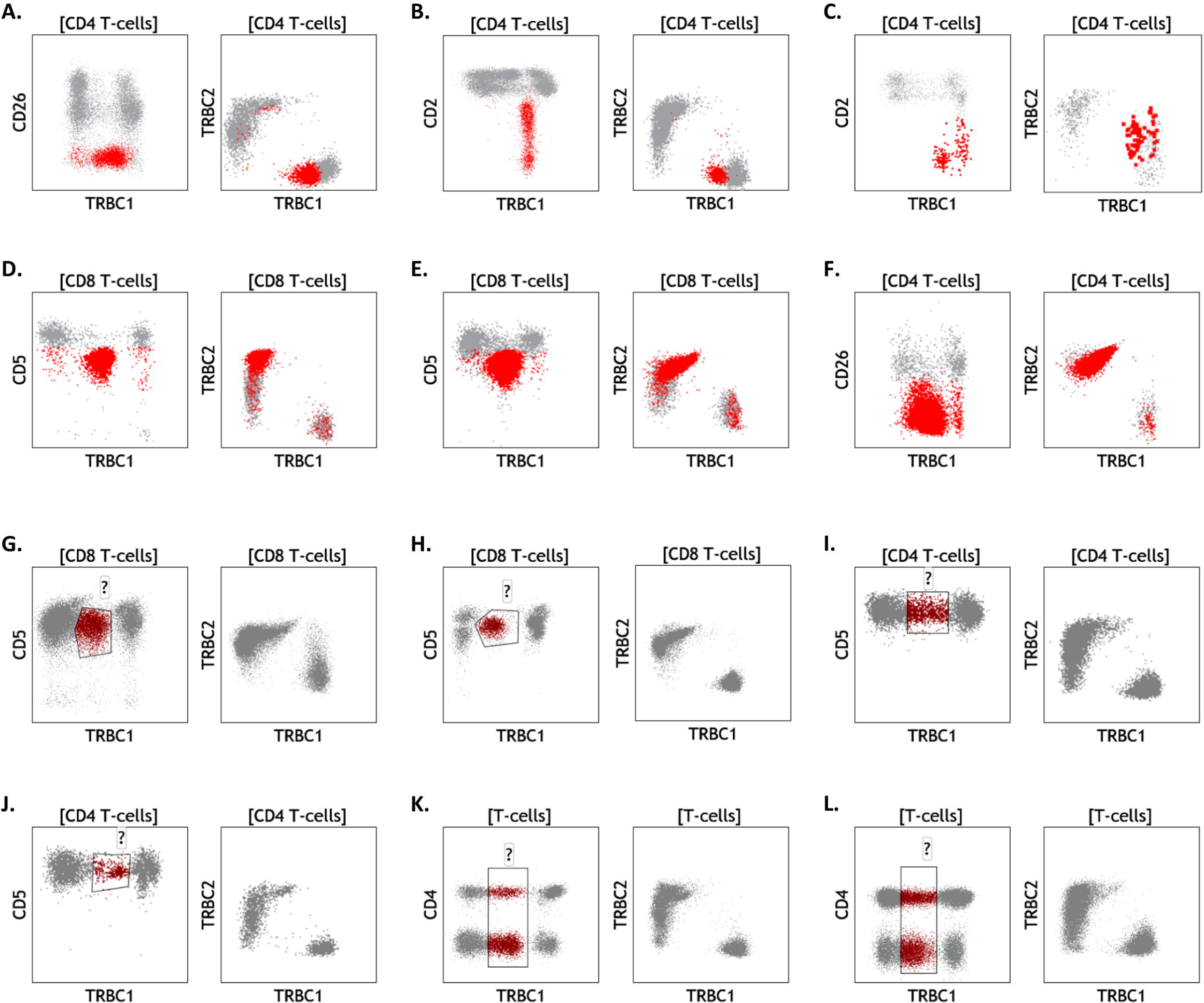
Dual T-cell receptor constant β chain (TRBC)1/TRBC2 assessment resolves common artifacts encountered with TRBC1-only staining. Specimens involved by cutaneous T-cell lymphoma (**A.**, **B.**, **C.** and **F.**), T-cell large granular lymphocytic leukemia (**D.** and **E.**), and samples from patients with no evidence of T-cell malignancy (**G.**-**L.**), were studied using TRBC1-only staining (left) or dual TRBC1/TRBC2 staining (right). Neoplastic cells (red) with spurious dim expression using TRBC1-only staining are clearly resolved as TRBC1-restricted (**A.-C.**) or TRBC2-restricted (**D.-F.**) neoplasms using dual TRBC1/TRBC2 staining. On benign samples, spurious dim-expressing subsets (maroon) detected with TRBC1-only staining on CD8^+^ T-cells (**G.** and **H.**), CD4^+^ T-cells (**I.** and **J.**) or both (**K.** and **L.**), are completely resolved with dual TRBC1/TRBC2 staining.

One of the caveats of testing for TRBC-restricted T-cell subsets by flow cytometry is the detection of common small T-CUS in a minority of patients without T-cell neoplasia, including healthy donors (**Figure 3F and 4B**). We have previously studied this phenomenon using our current diagnostic T-cell panel with TRBC1-only staining, and shown concurrent TCR-Vβ restriction and an immunophenotypic spectrum closely resembling that of T-LGLL.^15^ These findings are well in line with a breadth of literature describing the high prevalence of small clonal T-cell subsets, predominantly in the CD8^+^ T-cell compartment, which are believed to represent physiologic expansions of mostly effector/memory cytotoxic T-cells in response to EBV, CMV, resolved acute infections, neoplastic processes or other sources of antigen exposure.^30–37^ In contrast to commonly utilized molecular assays of T-cell clonality, flow cytometry with dual TRBC1 and TRBC2 staining provides useful information regarding the immunophenotype of the T-cell clone detected and its size relative to other T-cell or leukocyte subsets. These are valuable characteristics to establish or suggest a distinction between T-CUS and low-level involvement by a T-cell lymphoproliferative disorder based on expert interpretation and clinical correlation.^15^ Moreover, the ability to rapidly and routinely identify T-CUS using dual TRBC1 and TRBC2 stains should facilitate future studies to understand its biology and clinical significance; similar to how other laboratory advances have contributed to our understanding of small clonal B-cell, plasma cell and myeloid proliferations.

In conclusion, we describe a novel and simple strategy to detect clonal TCRαβ T-cells by flow cytometry using dual staining for TRBC1 and TRBC2 within a comprehensive single-tube T-cell panel. This approach can be easily adopted by clinical flow cytometry laboratories for routine analysis, and minimizes the need for a separate T-cell clonality assay. It also provides useful information about the TRBC isoform expressed by T-cell neoplasms for the selection of emerging CAR T-cell therapies targeting TRBC1 or TRBC2^13,14^. Our anti-TRBC2 antibody has recently been licensed to selected flow cytometry reagent manufacturers and will soon be commercially available.

## Data Availability

All data produced in the present work are contained in the manuscript

## AUTHOR CONTRIBUTIONS

PH, HO, MH and DJ designed the study of human samples; MP, PMM and MF designed and developed the anti-TRBC2 antibody; ZA and FTI preformed the non-clinical testing of the anti-TRBC2 antibody; MJW, GO and YP performed the flow cytometry experiments; PH and HO performed the flow cytometry analysis; PH, JO and JNS designed the machine learning analysis; PH and MF performed data analysis; PH wrote the manuscript; PMM, MP, DJ, MH, HO and MS edited the manuscript; HO, MS, MP, PMM and MH provided project support and expert knowledge.

