## Supplemental material for "Dual T-cell constant β chain (TRBC)1 and TRBC2 staining for the identification of T-cell neoplasms by flow cytometry"

**SUPPLEMENTAL FIGURE 1**

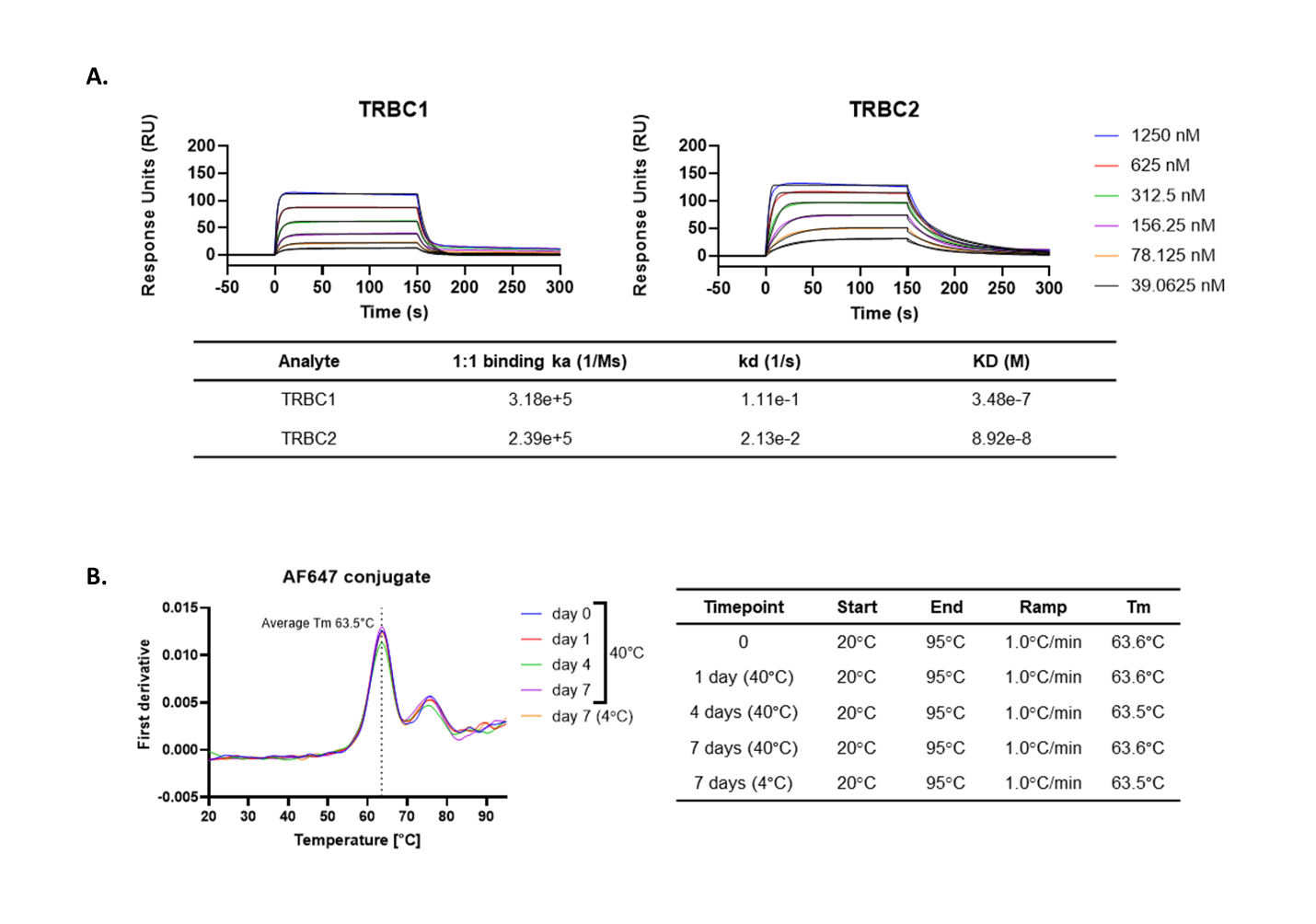

**Kinetic profile and thermal stability of a genetically engineered anti-TRBC2 antibody. A.** Kinetic profile of anti-TRBC2 antibody measured via Biacore T200 surface plasmon resonance system (Cytiva, Marlkborough, MA), against soluble TCRs carrying TRBC1 or TRBC2. Affinity for TRBC1 was measured at 348 nM, while affinity for TRBC2 was measured at 89 nM using a Langmuir 1:1 binding model. **B.** Thermal stability analysis of anti-TRBC2 AF647 conjugated antibody assessed via Prometheus NT.48 nanoDSF differential scanner fluorimeter (NanoTemper, München, Germany). The antibody was subjected to up to 7 days incubation at 40°C or 4°C. No significant shift of Tm was detected across the tested conditions. Anti-TRBC2 antibody, carrying murine IgG2a Fc, or unconjugated Jovi.1, carrying human IgG1 Fc, was included in the assay.

**SUPPLEMENTAL FIGURE 2**

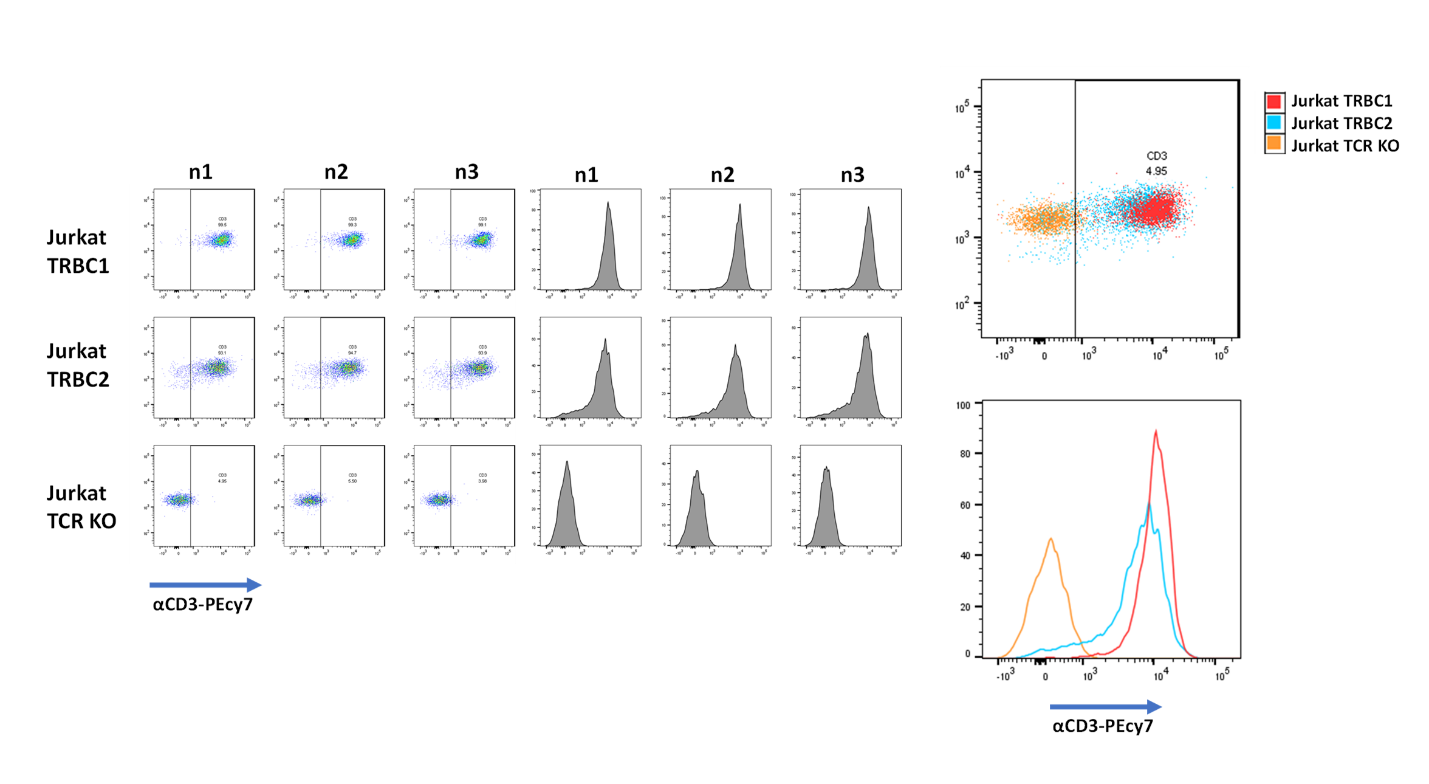

**Similar levels of CD3/TCR expression on TRBC1+ and TRBC2+ JURKAT cell lines.** Cells were stained by flow cytometry using anti‐CD3 PECy7 (Biolegend, San Diego, CA). *Left:* Representative flow cytometry dot plots and histograms on 3 batches of thawed cell line vials (n1, n2 and n3). *Right:* Overlayed dot plot and histogram comparing first batches only. Also shown are results of a Jurkat cell line where the CD3/TCR had been knocked out (Jurkat TCR KO).

**SUPPLEMENTAL FIGURE 3**

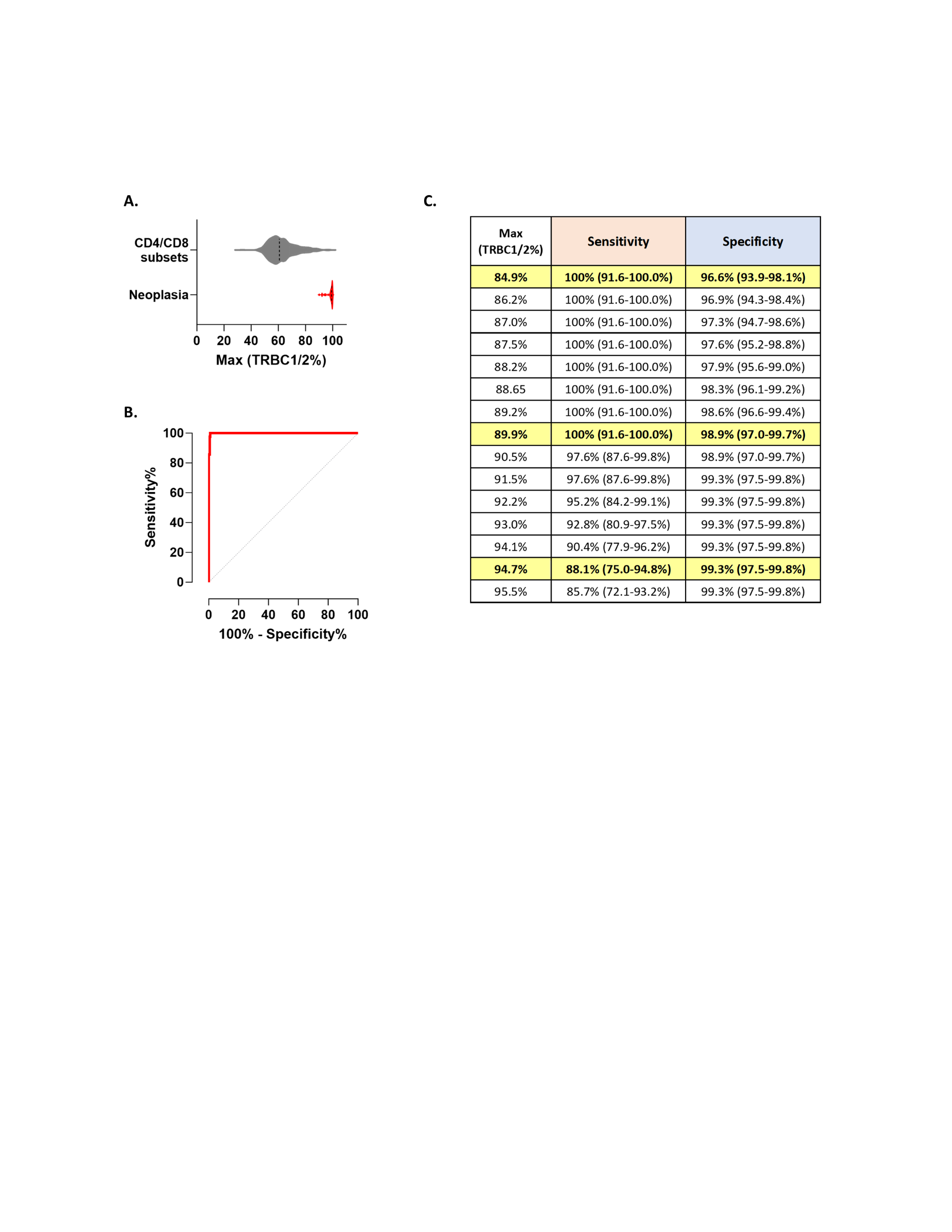

**Receiver operating curve analysis of TRBC-restriction thresholds for the diagnosis of T-cell neoplasia.** The maximum of %TRBC1^+^ and %TRBC2^+^ was studied as a single value to evaluate test performance. Gated tumor cells from 45 malignant cases (5 cases lacking intracellular TRBC expression excluded) were compared to gated CD4^+^/CD8^-^, CD8^+^/CD4^-^, CD4^+^/CD8^+^ and CD4^-^/CD8^-^ TCRαβ T-cell subsets (>200 events) from 109 samples without T-cell neoplasia (donors and benign samples studied with the T-cell panel). **A.** As expected, the median values were much higher for neoplastic (99.24%) compared to benign (60.75%) subsets. **B.** Receiver operator curve showing optimal performance, with an area under the curve of 0.999 (95% confidence interval: 0.9973 to 1.0), and a P significant of 0.0001. **C.** Sensitivity and specificity estimates for various thresholds, showing optimal performance for thresholds between 85% and 90%. The findings are consistent with our arbitrarily defined threshold of 85% based on extensive practice experience.

**SUPPLEMENTAL FIGURE 4**

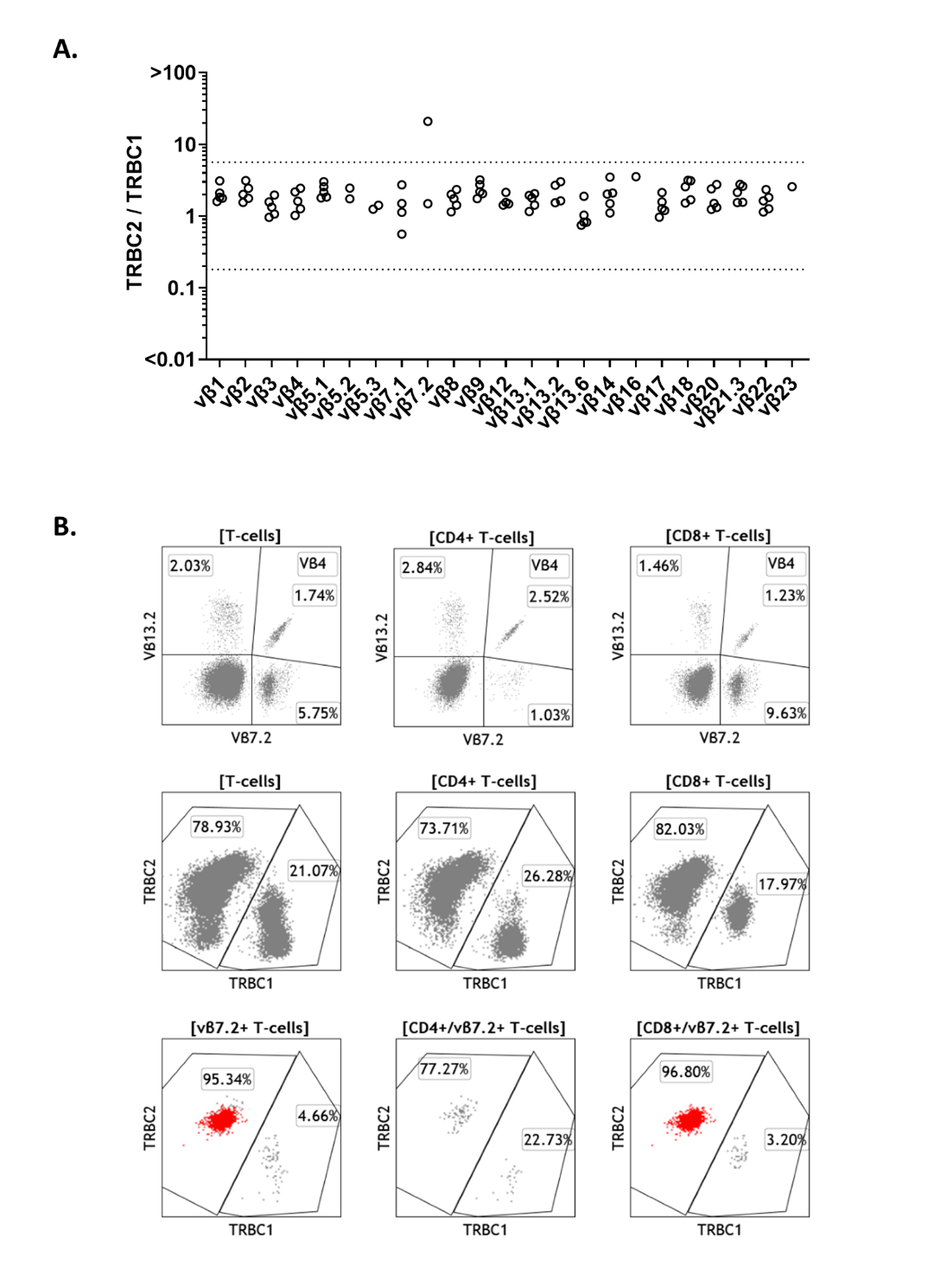

**Analysis of TRBC1 and TRBC2 expression on TCR-Vβ subsets from healthy donors.** Flow cytometric analysis on an outlier patient with a small TRBC2-restricted Vβ7.2 subset (see Figure 2D). No T-cell clonality was identified by Vβ analysis (*top*) or TRBC staining (*middle*) alone. However, combined analysis (*bottom*) revealed TRBC2-restriction on Vβ7.2-positive T-cells (bottom), corresponding to a small CD8-positive T-cell clone of uncertain significance (4.1% of lymphocytes).
